# Administration of tocilizumab to patients with high concentrations of IL-6 in the course of COVID-19 is associated with a better prognosis

**DOI:** 10.1101/2021.01.28.21249932

**Authors:** Robert Flisiak, Jerzy Jaroszewicz, Magdalena Rogalska, Tadeusz Łapiński, Aleksandra Berkan-Kawińska, Beata Bolewska, Magdalena Tudrujek-Zdunek, Dorota Kozielewicz, Marta Rorat, Piotr Leszczyński, Krzysztof Kłos, Justyna Kowalska, Paweł Pabjan, Anna Piekarska, Iwona Mozer-Lisewska, Krzysztof Tomasiewicz, Małgorzata Pawłowska, Krzysztof Simon, Joanna Polańska, Dorota Zarębska-Michaluk

## Abstract

**Background:** Despite the direct viral activity, the pathogenesis of coronavirus disease 2019 (COVID-19) includes an overproduction of cytokines including interleukin 6 (IL-6). Therefore tocilizumab (TCZ), a monoclonal antibody against IL-6 receptors, became considered as a possible therapeutic option.

**Methods:** Patients were selected from the SARSTer national database, which included 2332 individuals with COVID-19 and the current study included 825 adult patients with moderate to severe course. The retrospective analysis was performed in 170 patients treated with TCZ and 655 without this medication or any other anti-cytokine therapy. The end-points of treatment effectiveness were a rate of death, need for mechanical ventilation, and clinical improvement.

**Results:** Patients treated with TCZ were balanced compared to non-TCZ regarding gender, age, BMI, and prevalence of coexisting conditions. The reduced death rate was demonstrated in patients treated with TCZ and baseline IL-6 >100 pg/ml (hazard ratio [HR]: 0.27, 95% confidence interval [CI]:0.10-0.78), or those needing oxygen supplementations who worsened within 7 days of hospitalization (HR: 0.38, 95% CI:0.16-0.88). The best effectiveness of TCZ was achieved in patients with a combination of baseline IL-6>100 pg/ml and either SpO2≤90% (HR for death, mechanical ventilation, and clinical improvement after 21 or 28 days: 0.07, 0.14, 5.53, 5.18 respectively) or requiring oxygen supplementation (HR for death and clinical improvement after 21 or 28 days, 0.18, 2.66, 2.85 respectively).

**Conclusions:** Tocilizumab administered for COVID-19 in patients with a baseline concentration of IL-6>100 pg/ml is associated with reduced mortality and faster clinical improvement, particularly if there is a need for oxygen supplementation due to SpO2≤90%.

## Introduction

A novel coronavirus, named severe acute respiratory syndrome coronavirus 2 (SARS-CoV-2) was identified in December of 2019 and found to be responsible for an outbreak of respiratory tract infections discovered in Wuhan, China. The outbreak of the disease known as a coronavirus disease 2019 (COVID-19), was announced as a global pandemic by the World Health Organization (WHO) in March 2020. The search for effective therapy focused on repurposing of approved drugs with confirmed activity against other viruses, that included, for example, remdesivir (RDV), which was previously studied for the treatment of Ebola virus disease as well as SARS-CoV-1 and middle east respiratory syndrome (MERS) coronaviruses [1, 2]. Based on findings from phase III clinical trials and real-world experience study RDV received both American and European authorization [3, 4, 5]. Recommendations were also given to low□molecular□weight heparin and dexamethasone [6, 7]. However, the pathogenesis of COVID-19 is complicated and includes in addition to direct viral effect and coagulopathy, an overproduction of proinflammatory cytokines termed a cytokine storm, which is responsible for organ damage and is considered a major reason for death due to COVID-19 [8]. Unfortunately, standard anti-inflammatory treatments appear to be insufficient for controlling the cytokine storm. Concentrations of several proinflammatory cytokines, including interleukin (IL)-6, are substantially increased in patients with severe COVID-19 [9]. Higher concentrations of IL-6 was shown to be associated with faster progression of the disease and worse prognosis. Therefore, tocilizumab (TCZ), an inhibitor of the IL-6 receptors became considered as a possible therapeutic option [9, 10, 11]. Data from several studies have been contradictory mostly to difficulties in the selection of optimal population and finding the proper stage of the disease for administration [12, 13, 14]. Although the most recent randomized, double-blind, placebo-controlled trial by Stone et al. was not able to confirm the effectiveness of TCZ, authors did not exclude the possible benefit from interleukin-6 receptor blockade in some patient populations, because of wide confidence intervals for efficacy comparisons [14].

The purpose of the study is to search for the population of patients with severe COVID-19, which could obtain maximal benefit from the administration of tocilizumab, and identify the predictors of response to the treatment with this drug.

## Patients and Methods

Patients were selected from the SARSTer national database, which included 2332 patients treated between 1 March and 31 October 2020 in 30 Polish centers. This ongoing project, supported by the Polish Association of Epidemiologists and Infectiologists, is a national real-world experience study assessing treatment in patients with COVID-19. The decision about the treatment regimen was taken entirely by the treating physician concerning current knowledge and recommendations of the Polish Association of Epidemiologists and Infectiologists [15, 16, 17]. The SARSTer study was approved by the Ethical Committee of the Medical University of Białystok. If necessary the local bioethics committees approved experimental use of drugs in patients with COVID-19. The current study included 825 adult patients with moderate to severe course of the disease. Patients aged below 18 years, those with oxygen saturation >95%, or acute respiratory distress syndrome (ARDS) at baseline were excluded.

The retrospective analysis was carried out in 170 patients treated with tocilizumab (RoActemra, Roche Pharma AG) and 655 patients which did not receive this medication as well as any other monoclonal antibody directed against cytokine receptors. Tocilizumab was administered intravenously at 8 mg/kg (maximum dose: 800 mg) in a single dose (1□hour infusion). If no improvement was observed, the second dose was considered after 8 to 12 hours (administered in 42% patients) according to the national recommendations [15, 16, 17]. Data were entered retrospectively and submitted online by a web-based platform operated by Tiba sp. z o.o. Parameters collected at baseline included age, gender, body mass index (BMI), coexisting conditions, other medication-related to COVID-19, clinical status at admission, and adverse events. Baseline clinical status at hospital admission was classified according to oxygen saturation (SpO_2_) 91-95%, or SpO_2_ ≤90% as well as based on the score on an ordinal scale.

The end-points of treatment effectiveness were a rate of death, need for mechanical ventilation, and clinical improvement in the ordinal scale based on WHO recommendations modified to fit the specificity of the national health care system. Clinical improvement was defined as at least a 2-point decrease from baseline to 14, 21, and 28 days of hospitalization. The ordinal scale was scored as follows: 1. unhospitalized, no activity restrictions; 2. unhospitalized, no activity restrictions and/or requiring oxygen supplementation at home; 3. hospitalized, does not require oxygen supplementation and does not require medical care; 4. hospitalized, requiring no oxygen supplementation, but requiring medical care; 5. hospitalized, requiring normal oxygen supplementation; 6. hospitalized, on non-invasive ventilation with high-flow oxygen equipment; 7. hospitalized, for invasive mechanical ventilation or extracorporeal membrane oxygenation (ECMO); 8. death.

To identify possible predictors of response to the treatment with TCZ we compared rates of achieved end-points in patients receiving versus not receiving TCZ. Following baseline, predictors were included: age above 70 years, the need for oxygen high flow (ordinal scale 6 points) at baseline, clinical worsening during 7 days of hospitalization in patients with regular oxygen supplementation at baseline (5 points in original scale), SpO2<90% at baseline and several laboratory measures at baseline, such as IL-6>100 pg/ml, C-reactive protein (CRP)>200 mg/l, neutrophils >7500 /μl, lymphocytes >1200 /μl, D-dimer >1000 μg/l, and procalcitonin >0.1 ng/ml.

### Statistical analysis

The results are expressed as mean ± standard deviation (SD) or n (%) and odds ratios with 95% confidence intervals. P values of <0.05 were considered to be statistically significant. The significance of difference was calculated by Fisher’s exact test for nominal variables and by Mann-Whitney U and Kruskal-Wallis ANOVA for continuous variables. Survival analyses were performed by Log-rank (Mantel-Cox) Test and depicted as Kaplan-Meier plots. Univariable comparisons were calculated by GraphPad Prism 5.1 (GraphPad Software, Inc., La Jolla, CA).

## Results

Among 825 patients included in the study 170 received therapy with TCZ and 655 did not receive TCZ. As shown in Table 1 groups were balanced regarding gender, age, and BMI, but there was a predominance of males in both arms. Patients treated with TCZ more frequently demonstrated a course of the disease with SpO_2_≤90% at admission to the hospital (65.9%) compared to those without TCZ (37.7%). Moreover, patients treated with TCZ more often required normal or high flow oxygen supplementation (93.6%) compared to the non-TCZ group (76.8%). The prevalence of coexisting conditions was similar in both groups, but patients treated with TCZ more frequently received other medication-related to COVID-19 (Table 1).

**Table 1.** Baseline demographic and clinical characteristics of included patients.

| Characteristic | All patients<br>N=825 | Tocilizumab<br>N=170 | No<br>Tocilizumab<br>N=655 |
| --- | --- | --- | --- |
| **Age** |  |  |  |
| Mean (SD) | 63.1 (15.1) | 63.2 (13.8) | 63.0 (15.4) |
| >70 years (%) | 267 (32.4) | 53 (31.2) | 214 (32.7) |
| **Gender** |  |  |  |
| Female, n (%) | 337 (40.8) | 60 (35.3) | 277 (42.3) |
| Male, n (%) | 488 (59.2) | 110 (64.7) | 378 (57.7) |
| **Body mass index, mean (SD)** | 28.8 (4.9) | 29.7 (4.8) | 28.5 (5.0) |
| **Disease severity at the baseline, n (%)** |  |  |  |
| oxygen saturation 91-95% | 466 (56.5) | 58 (34.1) | 408 (62.3) |
| oxygen saturation ≤90% | 359 (43.5) | 112 (65.9) | 247 (37.7) |
| **Score on ordinal scale, n (%)** |  |  |  |
| 4. hospitalized, requiring no oxygen supplementation, but requiring medical care | 163 (19.8) | 11 (6.5) | 152 (23.2) |
| 5. hospitalized, requiring normal oxygen supplementation | 615 (74.5) | 147 (86.5) | 468 (71.5) |
| 6. hospitalized, on non-invasive ventilation with high-flow oxygen equipment | 47 (5.7) | 12 (7.1) | 35 (5.3) |
| **Coexisting conditions, n (%)** | 638 (77.3) | 131 (77.1) | 507 (77.4) |
| **Other medication-related to COVID-19, n (%)** |  |  |  |
| Remdesivir | 284 (34.4) | 67 (39.4) | 217 (33.1) |
| Dexamethason | 272 (33.0) | 83 (48.8) | 189 (28.9) |
| Convalescent plasma | 103 (12.5) | 29 (17.1) | 74 (11.3) |
| Low molecular weight heparin | 713 (86.5) | 170 (100.0) | 543 (82.9) |

As shown in Table 2 the rate of clinical improvement after 21 and 28 days was significantly higher in patients which did not receive TCZ. However, a statistically significant decrease in death rate was demonstrated in patients treated with TCZ and baseline IL-6 exceeding 100 pg/ml or those needing oxygen supplementation at baseline who worsened within the initial 7 days of hospitalization (Table 2, Figure 1). As shown with the Kaplan-Meier analysis there were no significant differences between TCZ and non-TCZ arms when the analysis was carried out in all patients or those with baseline IL-6 concentration below 100 pg/ml. Further analysis included the correlation between IL-6 concentration and several possible clinical and laboratory indices associated with the course of the disease. Among patients with baseline SpO_2_≤90%, which are potential TCZ recipients significant correlation was demonstrated between serum concentrations of IL-6 and SpO_2_, levels of C-reactive protein, procalcitonin D-dimers, as well as white blood cell and neutrophil counts (Table 3).

**Table 2.**
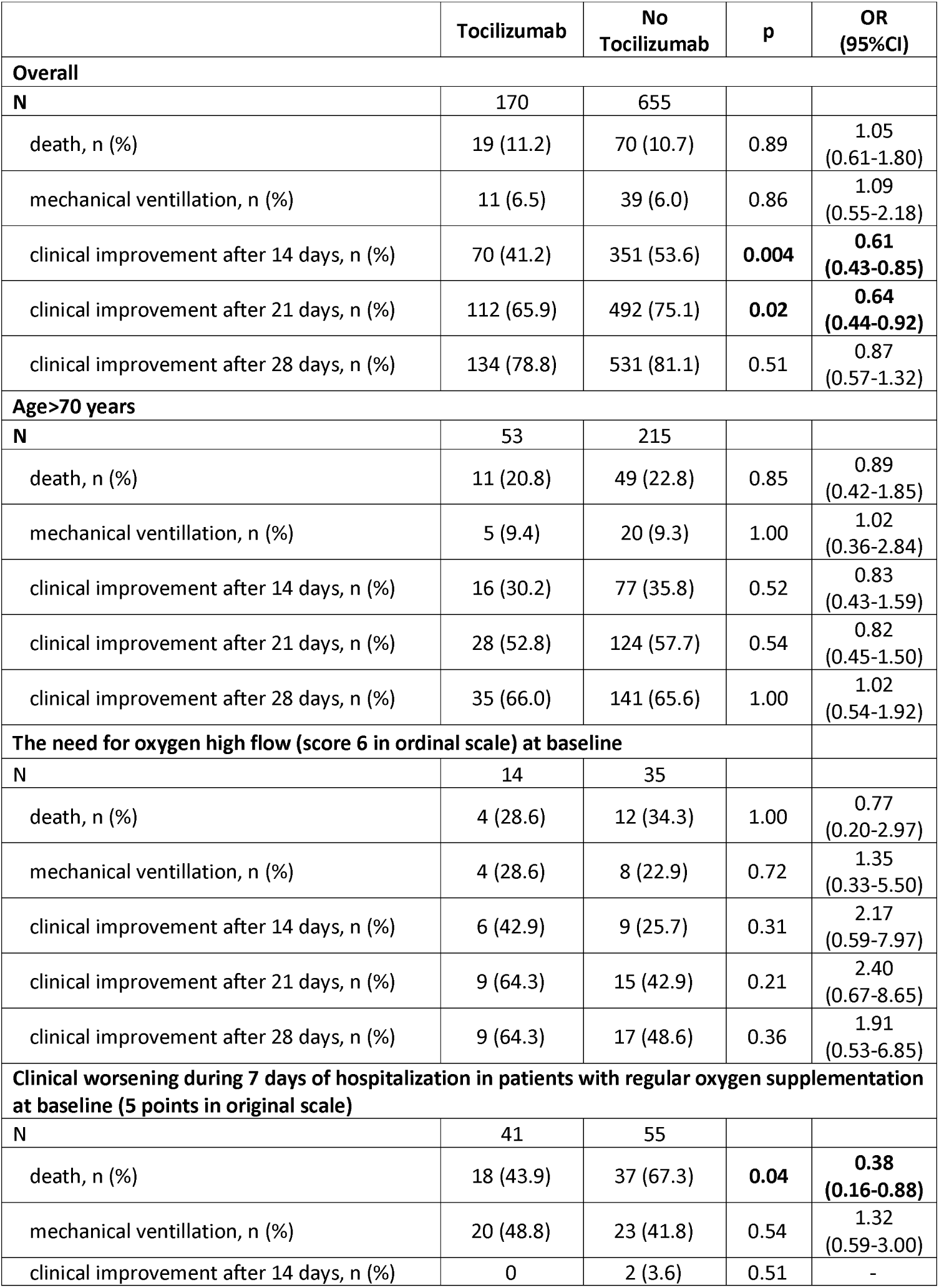
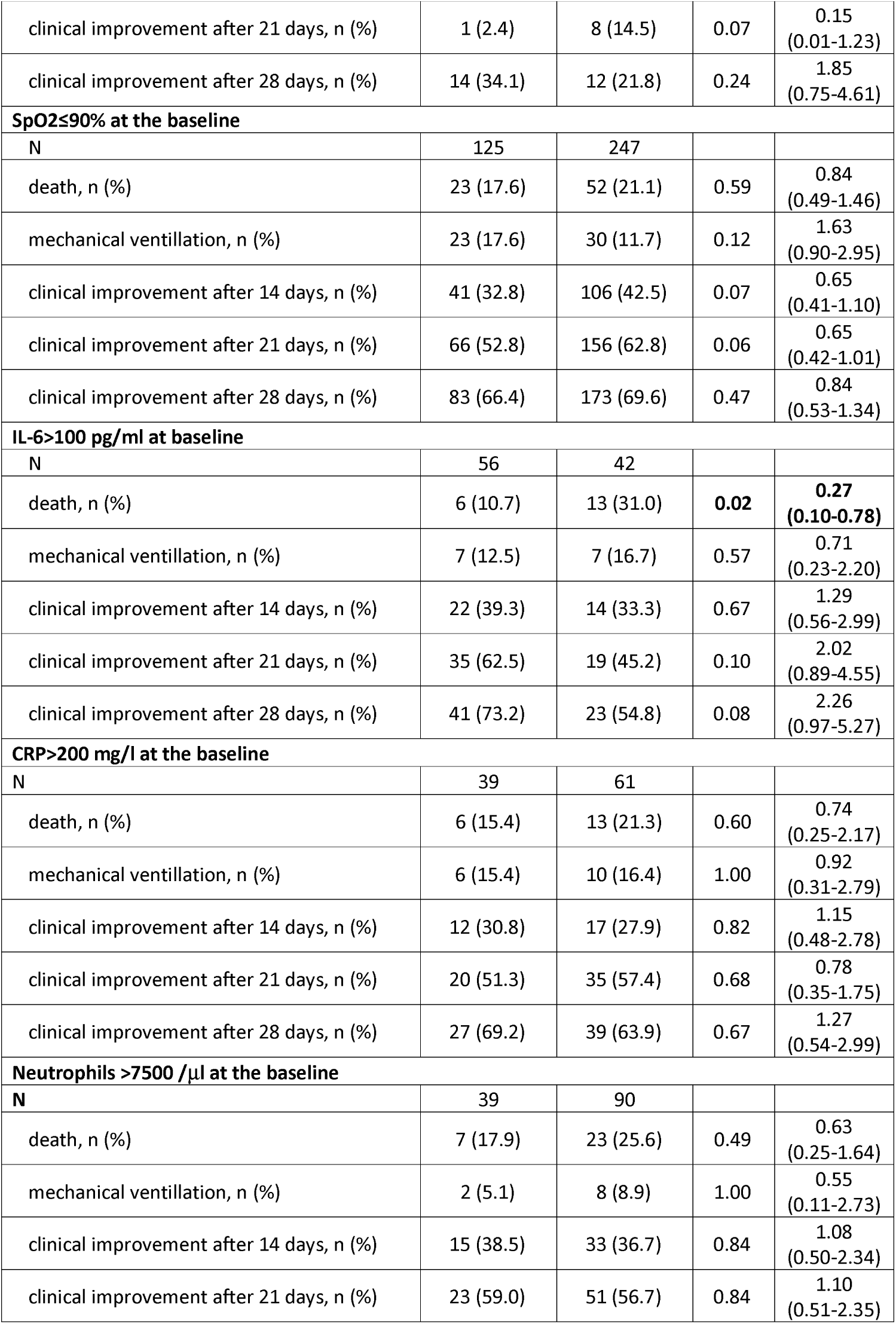
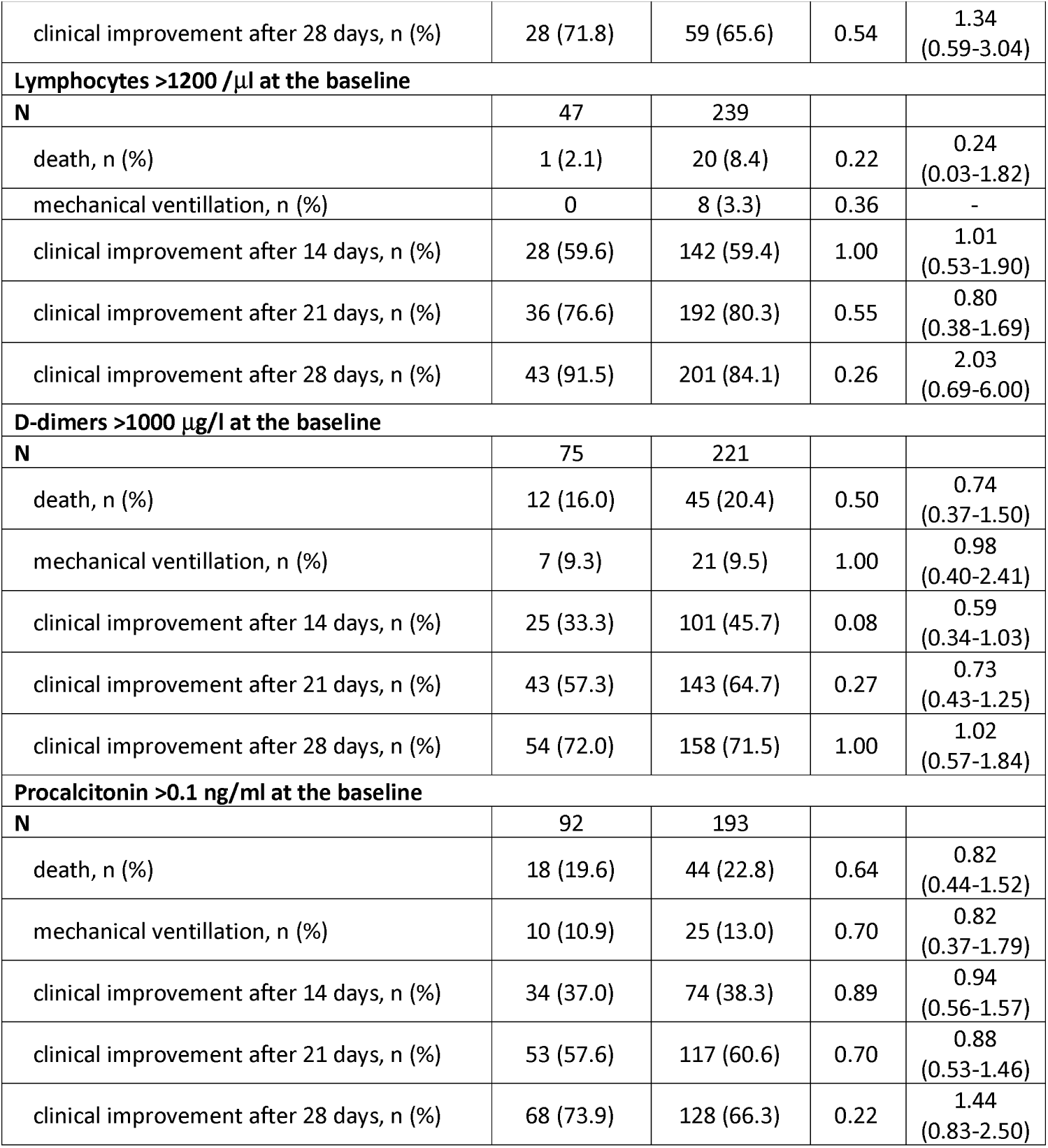
Tocilizumab effect on rates of death need for mechanical ventilation and clinical improvement depending on possible outcomes predictors.

|  | **Tocilizumab** | **No**<br>**Tocilizumab** | **p** | **OR**<br>**(95%CI)** |
| --- | --- | --- | --- | --- |
| **Overall** |  |  |  |  |
| **N** | 170 | 655 |  |  |
| death, n (%) | 19 (11.2) | 70 (10.7) | 0.89 | 1.05<br>(0.61-1.80) |
| mechanical ventilation, n (%) | 11 (6.5) | 39 (6.0) | 0.86 | 1.09<br>(0.55-2.18) |
| clinical improvement after 14 days, n (%) | 70 (41.2) | 351 (53.6) | **0.004** | **0.61**<br>**(0.43-0.85)** |
| clinical improvement after 21 days, n (%) | 112 (65.9) | 492 (75.1) | **0.02** | **0.64**<br>**(0.44-0.92)** |
| clinical improvement after 28 days, n (%) | 134 (78.8) | 531 (81.1) | 0.51 | 0.87<br>(0.57-1.32) |
| **Age>70 years** |  |  |  |  |
| **N** | 53 | 215 |  |  |
| death, n (%) | 11 (20.8) | 49 (22.8) | 0.85 | 0.89<br>(0.42-1.85) |
| mechanical ventilation, n (%) | 5 (9.4) | 20 (9.3) | 1.00 | 1.02<br>(0.36-2.84) |
| clinical improvement after 14 days, n (%) | 16 (30.2) | 77 (35.8) | 0.52 | 0.83<br>(0.43-1.59) |
| clinical improvement after 21 days, n (%) | 28 (52.8) | 124 (57.7) | 0.54 | 0.82<br>(0.45-1.50) |
| clinical improvement after 28 days, n (%) | 35 (66.0) | 141 (65.6) | 1.00 | 1.02<br>(0.54-1.92) |
| **The need for oxygen high flow (score 6 in ordinal scale) at baseline** |  |  |  |  |
| **N** | 14 | 35 |  |  |
| death, n (%) | 4 (28.6) | 12 (34.3) | 1.00 | 0.77<br>(0.20-2.97) |
| mechanical ventilation, n (%) | 4 (28.6) | 8 (22.9) | 0.72 | 1.35<br>(0.33-5.50) |
| clinical improvement after 14 days, n (%) | 6 (42.9) | 9 (25.7) | 0.31 | 2.17<br>(0.59-7.97) |
| clinical improvement after 21 days, n (%) | 9 (64.3) | 15 (42.9) | 0.21 | 2.40<br>(0.67-8.65) |
| clinical improvement after 28 days, n (%) | 9 (64.3) | 17 (48.6) | 0.36 | 1.91<br>(0.53-6.85) |
| **Clinical worsening during 7 days of hospitalization in patients with regular oxygen supplementation at baseline (5 points in original scale)** |  |  |  |  |
| **N** | 41 | 55 |  |  |
| death, n (%) | 18 (43.9) | 37 (67.3) | **0.04** | **0.38**<br>**(0.16-0.88)** |
| mechanical ventilation, n (%) | 20 (48.8) | 23 (41.8) | 0.54 | 1.32<br>(0.59-3.00) |
| clinical improvement after 14 days, n (%) | 0 | 2 (3.6) | 0.51 | - |
| clinical improvement after 21 days, n (%) | 1 (2.4) | 8 (14.5) | 0.07 | 0.15<br>(0.01-1.23) |
| clinical improvement after 28 days, n (%) | 14 (34.1) | 12 (21.8) | 0.24 | 1.85<br>(0.75-4.61) |
| **SpO2≤90% at the baseline** |  |  |  |  |
| N | 125 | 247 |  |  |
| death, n (%) | 23 (17.6) | 52 (21.1) | 0.59 | 0.84<br>(0.49-1.46) |
| mechanical ventilation, n (%) | 23 (17.6) | 30 (11.7) | 0.12 | 1.63<br>(0.90-2.95) |
| clinical improvement after 14 days, n (%) | 41 (32.8) | 106 (42.5) | 0.07 | 0.65<br>(0.41-1.10) |
| clinical improvement after 21 days, n (%) | 66 (52.8) | 156 (62.8) | 0.06 | 0.65<br>(0.42-1.01) |
| clinical improvement after 28 days, n (%) | 83 (66.4) | 173 (69.6) | 0.47 | 0.84<br>(0.53-1.34) |
| **IL-6>100 pg/ml at baseline** |  |  |  |  |
| N | 56 | 42 |  |  |
| death, n (%) | 6 (10.7) | 13 (31.0) | **0.02** | **0.27**<br>**(0.10-0.78)** |
| mechanical ventilation, n (%) | 7 (12.5) | 7 (16.7) | 0.57 | 0.71<br>(0.23-2.20) |
| clinical improvement after 14 days, n (%) | 22 (39.3) | 14 (33.3) | 0.67 | 1.29<br>(0.56-2.99) |
| clinical improvement after 21 days, n (%) | 35 (62.5) | 19 (45.2) | 0.10 | 2.02<br>(0.89-4.55) |
| clinical improvement after 28 days, n (%) | 41 (73.2) | 23 (54.8) | 0.08 | 2.26<br>(0.97-5.27) |
| **CRP>200 mg/l at the baseline** |  |  |  |  |
| N | 39 | 61 |  |  |
| death, n (%) | 6 (15.4) | 13 (21.3) | 0.60 | 0.74<br>(0.25-2.17) |
| mechanical ventilation, n (%) | 6 (15.4) | 10 (16.4) | 1.00 | 0.92<br>(0.31-2.79) |
| clinical improvement after 14 days, n (%) | 12 (30.8) | 17 (27.9) | 0.82 | 1.15<br>(0.48-2.78) |
| clinical improvement after 21 days, n (%) | 20 (51.3) | 35 (57.4) | 0.68 | 0.78<br>(0.35-1.75) |
| clinical improvement after 28 days, n (%) | 27 (69.2) | 39 (63.9) | 0.67 | 1.27<br>(0.54-2.99) |
| **Neutrophils >7500 /μl at the baseline** |  |  |  |  |
| N | 39 | 90 |  |  |
| death, n (%) | 7 (17.9) | 23 (25.6) | 0.49 | 0.63<br>(0.25-1.64) |
| mechanical ventilation, n (%) | 2 (5.1) | 8 (8.9) | 1.00 | 0.55<br>(0.11-2.73) |
| clinical improvement after 14 days, n (%) | 15 (38.5) | 33 (36.7) | 0.84 | 1.08<br>(0.50-2.34) |
| clinical improvement after 21 days, n (%) | 23 (59.0) | 51 (56.7) | 0.84 | 1.10<br>(0.51-2.35) |
| clinical improvement after 28 days, n (%) | 28 (71.8) | 59 (65.6) | 0.54 | 1.34<br>(0.59-3.04) |
| **Lymphocytes >1200 /μl at the baseline** |  |  |  |  |
| **N** | 47 | 239 |  |  |
| death, n (%) | 1 (2.1) | 20 (8.4) | 0.22 | 0.24<br>(0.03-1.82) |
| mechanical ventilation, n (%) | 0 | 8 (3.3) | 0.36 | - |
| clinical improvement after 14 days, n (%) | 28 (59.6) | 142 (59.4) | 1.00 | 1.01<br>(0.53-1.90) |
| clinical improvement after 21 days, n (%) | 36 (76.6) | 192 (80.3) | 0.55 | 0.80<br>(0.38-1.69) |
| clinical improvement after 28 days, n (%) | 43 (91.5) | 201 (84.1) | 0.26 | 2.03<br>(0.69-6.00) |
| **D-dimers >1000 μg/l at the baseline** |  |  |  |  |
| **N** | 75 | 221 |  |  |
| death, n (%) | 12 (16.0) | 45 (20.4) | 0.50 | 0.74<br>(0.37-1.50) |
| mechanical ventilation, n (%) | 7 (9.3) | 21 (9.5) | 1.00 | 0.98<br>(0.40-2.41) |
| clinical improvement after 14 days, n (%) | 25 (33.3) | 101 (45.7) | 0.08 | 0.59<br>(0.34-1.03) |
| clinical improvement after 21 days, n (%) | 43 (57.3) | 143 (64.7) | 0.27 | 0.73<br>(0.43-1.25) |
| clinical improvement after 28 days, n (%) | 54 (72.0) | 158 (71.5) | 1.00 | 1.02<br>(0.57-1.84) |
| **Procalcitonin >0.1 ng/ml at the baseline** |  |  |  |  |
| **N** | 92 | 193 |  |  |
| death, n (%) | 18 (19.6) | 44 (22.8) | 0.64 | 0.82<br>(0.44-1.52) |
| mechanical ventilation, n (%) | 10 (10.9) | 25 (13.0) | 0.70 | 0.82<br>(0.37-1.79) |
| clinical improvement after 14 days, n (%) | 34 (37.0) | 74 (38.3) | 0.89 | 0.94<br>(0.56-1.57) |
| clinical improvement after 21 days, n (%) | 53 (57.6) | 117 (60.6) | 0.70 | 0.88<br>(0.53-1.46) |
| clinical improvement after 28 days, n (%) | 68 (73.9) | 128 (66.3) | 0.22 | 1.44<br>(0.83-2.50) |

**Table 3.** Correlations between baseline serum IL-6 vs selected clinical and laboratory indices.

| IL-6 versus | All patients<br>(n=825) |  | SpO<sub>2</sub> ≤90%<br>(n=372) |  | SpO<sub>2</sub> 91-95%<br>(n=453) |  |
| --- | --- | --- | --- | --- | --- | --- |
|  | R | p | R | p | R | P |
| Age | 0.15 | <0.001 | 0.07 | 0.32 | 0.18 | 0.002 |
| BMI | 0.01 | 0.86 | -0.03 | 0.63 | 0.01 | 0.89 |
| SpO<sub>2</sub> | -0.33 | <0.001 | -0.19 | 0.003 | -0.31 | <0.001 |
| CRP | 0.58 | <0.001 | 0.44 | <0.001 | 0.68 | <0.001 |
| Procalcitonin | 0.40 | <0.001 | 0.37 | <0.001 | 0.36 | <0.001 |
| WBC | 0.26 | <0.001 | 0.22 | <0.001 | 0.25 | <0.001 |
| Lymphocytes | -0.21 | <0.001 | -0.05 | 0.42 | -0.28 | <0.001 |
| Neutrophils | 0.32 | <0.001 | 0.23 | <0.001 | 0.37 | <0.001 |
| Platelets | -0.10 | 0.04 | -0.10 | 0.14 | -0.13 | 0.03 |
| D-dimers | 0.28 | <0.001 | 0.20 | 0.003 | 0.27 | <0.001 |
| ALT | 0.13 | 0.03 | 0.10 | 0.13 | 0.10 | 0.09 |

**Figure 1.**
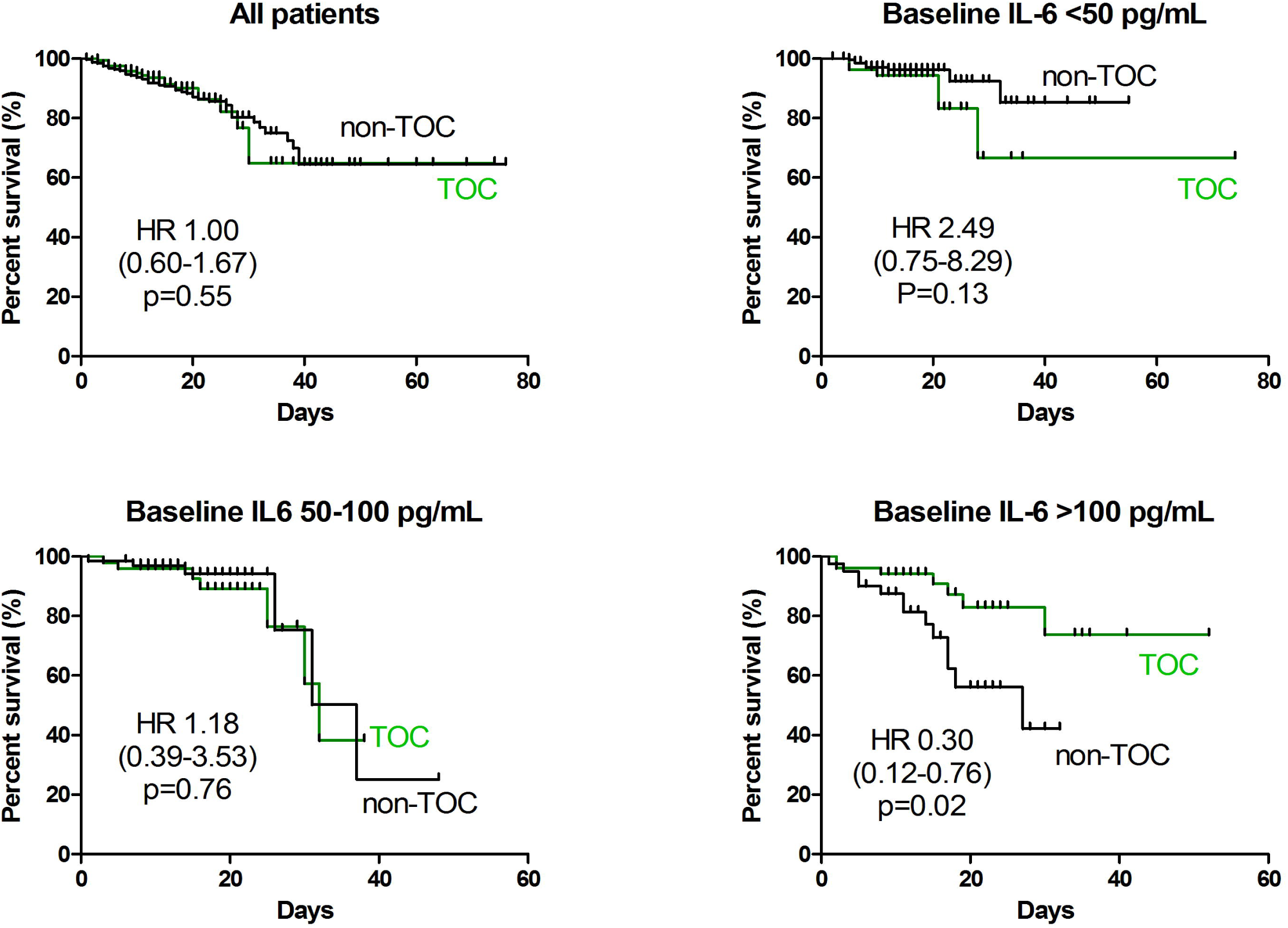
Kaplan-Meier graphs demonstrating the effect of tocilizumab versus no tocilizumab administration on patients survival depending on baseline serum concentration of interleukin 6.

We also analyzed a combination of several measures. As shown in table 4, the best effectiveness of TCZ administration can be achieved in patients with serum IL-6>100 pg/ml and either SpO_2_≤90% or requiring normal or high-flow oxygen supplementation. Statistically, significant effectiveness was achieved regarding the risk of death, the need for mechanical ventilation, as well as clinical improvement after 21 and 28 days (Table 4). Significantly better survival among such patients treated with TCZ was also demonstrated with a Kaplan-Meier analysis (Figure 2).

**Table 4.**
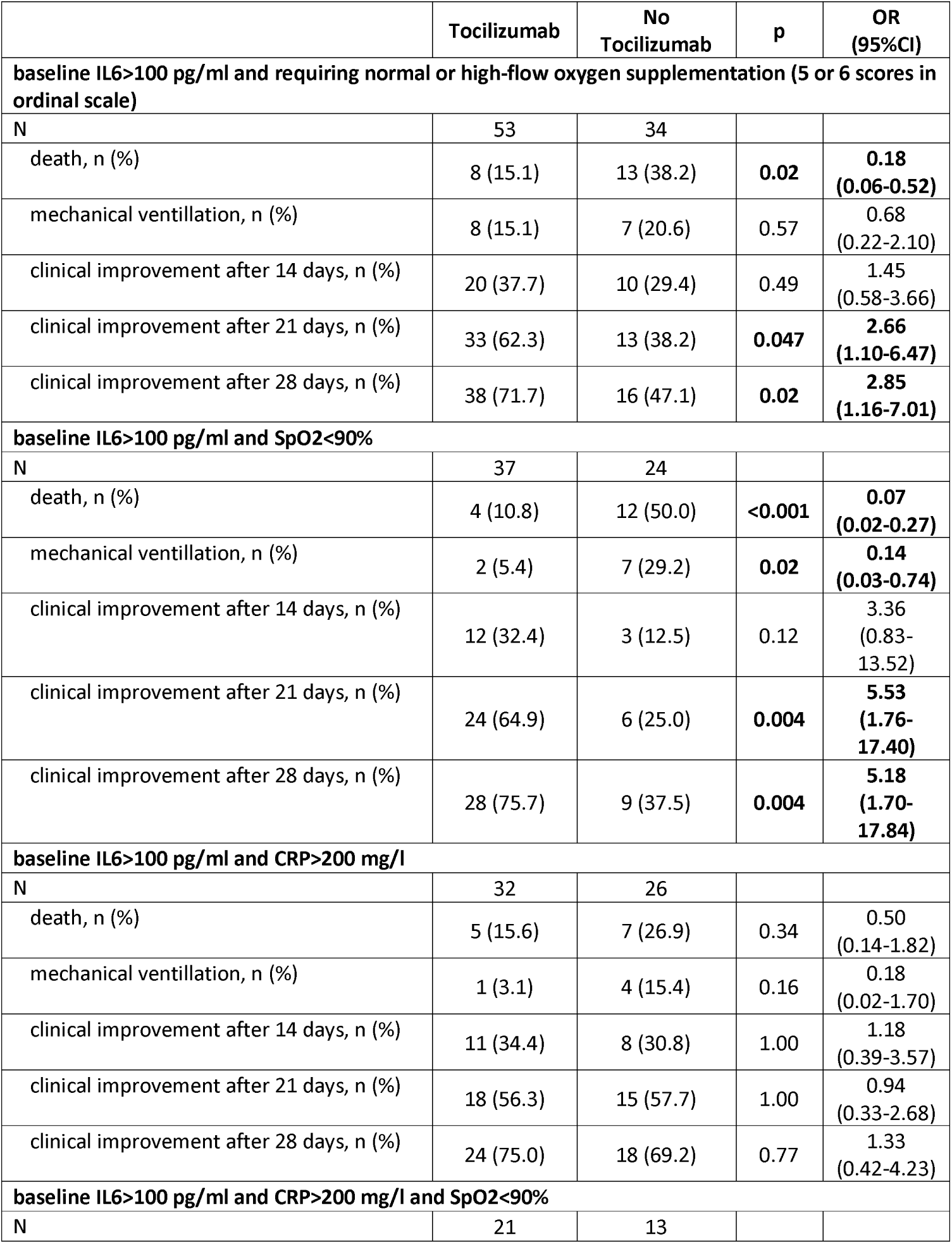
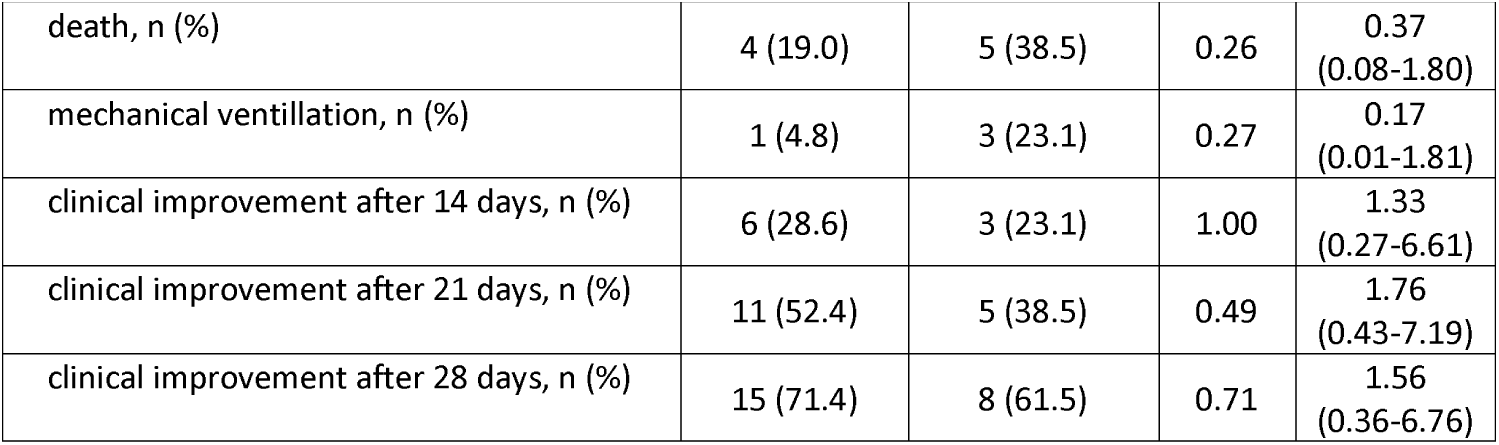
Tocilizumab effect on rates of death need for mechanical ventilation, and clinical improvement depending on combinations of possible outcomes predictors.

|  | **Tocilizumab** | **No**<br>**Tocilizumab** | **p** | **OR**<br>**(95%CI)** |
| --- | --- | --- | --- | --- |
| **baseline IL6>100 pg/ml and requiring normal or high-flow oxygen supplementation (5 or 6 scores in ordinal scale)** |  |  |  |  |
| **N** | 53 | 34 |  |  |
| death, n (%) | 8 (15.1) | 13 (38.2) | **0.02** | **0.18**<br>**(0.06-0.52)** |
| mechanical ventilation, n (%) | 8 (15.1) | 7 (20.6) | 0.57 | 0.68<br>(0.22-2.10) |
| clinical improvement after 14 days, n (%) | 20 (37.7) | 10 (29.4) | 0.49 | 1.45<br>(0.58-3.66) |
| clinical improvement after 21 days, n (%) | 33 (62.3) | 13 (38.2) | **0.047** | **2.66**<br>**(1.10-6.47)** |
| clinical improvement after 28 days, n (%) | 38 (71.7) | 16 (47.1) | **0.02** | **2.85**<br>**(1.16-7.01)** |
| **baseline IL6>100 pg/ml and SpO2<90%** |  |  |  |  |
| **N** | 37 | 24 |  |  |
| death, n (%) | 4 (10.8) | 12 (50.0) | **<0.001** | **0.07**<br>**(0.02-0.27)** |
| mechanical ventilation, n (%) | 2 (5.4) | 7 (29.2) | **0.02** | **0.14**<br>**(0.03-0.74)** |
| clinical improvement after 14 days, n (%) | 12 (32.4) | 3 (12.5) | 0.12 | 3.36<br>(0.83-13.52) |
| clinical improvement after 21 days, n (%) | 24 (64.9) | 6 (25.0) | **0.004** | **5.53**<br>**(1.76-17.40)** |
| clinical improvement after 28 days, n (%) | 28 (75.7) | 9 (37.5) | **0.004** | **5.18**<br>**(1.70-17.84)** |
| **baseline IL6>100 pg/ml and CRP>200 mg/l** |  |  |  |  |
| **N** | 32 | 26 |  |  |
| death, n (%) | 5 (15.6) | 7 (26.9) | 0.34 | 0.50<br>(0.14-1.82) |
| mechanical ventilation, n (%) | 1 (3.1) | 4 (15.4) | 0.16 | 0.18<br>(0.02-1.70) |
| clinical improvement after 14 days, n (%) | 11 (34.4) | 8 (30.8) | 1.00 | 1.18<br>(0.39-3.57) |
| clinical improvement after 21 days, n (%) | 18 (56.3) | 15 (57.7) | 1.00 | 0.94<br>(0.33-2.68) |
| clinical improvement after 28 days, n (%) | 24 (75.0) | 18 (69.2) | 0.77 | 1.33<br>(0.42-4.23) |
| **baseline IL6>100 pg/ml and CRP>200 mg/l and SpO2<90%** |  |  |  |  |
| **N** | 21 | 13 |  |  |
| death, n (%) | 4 (19.0) | 5 (38.5) | 0.26 | 0.37<br>(0.08-1.80) |
| mechanical ventilation, n (%) | 1 (4.8) | 3 (23.1) | 0.27 | 0.17<br>(0.01-1.81) |
| clinical improvement after 14 days, n (%) | 6 (28.6) | 3 (23.1) | 1.00 | 1.33<br>(0.27-6.61) |
| clinical improvement after 21 days, n (%) | 11 (52.4) | 5 (38.5) | 0.49 | 1.76<br>(0.43-7.19) |
| clinical improvement after 28 days, n (%) | 15 (71.4) | 8 (61.5) | 0.71 | 1.56<br>(0.36-6.76) |

**Figure 2.**
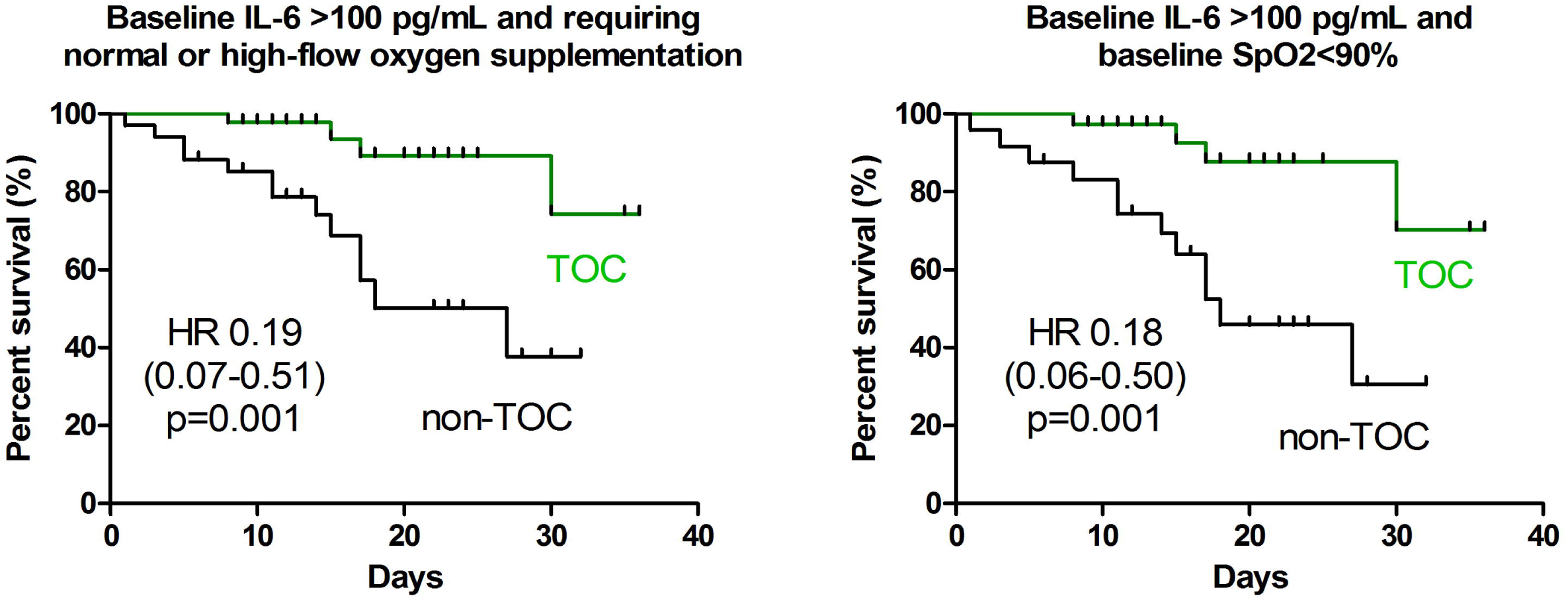
Kaplan-Meier graphs demonstrating the effect of tocilizumab versus no tocilizumab administration on patients survival depending on baseline serum concentration of interleukin 6.

Adverse events related to therapy were infrequent and reported in 22% of patients treated with TCZ. The most frequent was an elevation of aminotransferases activities (10%), which was usually associated with remdesivir. Less frequent were diarrhea (5%) - mostly in patients receiving lopinavir/ritonavir, and prolonged QT interval (3%) related to chloroquine.

## Discussion

Uncontrolled immune activation with high-level release of various pro-inflammatory cytokines is a hallmark of the lung, but also multiorgan damage, during later phases of COVID-19. It is usually termed as a “cytokine storm” and observed mainly during the second and third week of symptomatic disease in patients with severely impaired oxygen saturation [18]. Unsurprisingly, anti-cytokine agents including anti-IL-1R and anti-IL6R antagonists were among important candidates in the therapy of later stages of COVID-19. One of the rationales was a good suppressive effect of tocilizumab in cytokine release syndrome during CAR T-cells therapy [19]. Regardless of the pathophysiological link various randomized and observational studies of TCZ, while suggesting some benefit, did not bring clear evidence supporting its use in COVID-19. This may reflect the unique features of individual immune responses to pathogens, as well as the necessity of a complex and personalized approach in prescribing and timing immunomodulatory treatment. Proposing clinical trial protocol taking such diversity into account proves to be challenging thus personalized medicine relies on observational research and real-life experiences.

Results of our real-world evidence study could potentially explain the lack of effectiveness of TCZ observed in other studies, but also provide information on the optimal use of this agent. In our cohort, similarly to the first two randomized controlled trials (RCT), TCZ did not decrease overall mortality in hospitalized patients with COVID-19. In the first by Stone JH et al. [14], 83% of 243 COVID-19 subjects requiring oxygen supplementation but not mechanical ventilation, were randomized to TCZ (8mg/kg, single dose) or placebo. In this study, the odd ratio (HR) for death was 0.83, which was not significant, but with broad 95% confidence intervals (0.38 to 1.81, P=0.64) suggesting heterogeneity of probable association depending on other clinical variables not found in the publication. In another RCT by Hermine et al [12] including 131 patients with COVID-19 pneumonia requiring oxygen supplementation but not mechanically ventilated, 64 were randomized to TCZ (8 mg/kg, twice) or placebo. Likewise, the adjusted HR for 28-day mortality was 0.92 (90%CI 0.33-2.53). On the other hand, on day 14 in TCZ-group, 12% fewer patients needed non-invasive or mechanical ventilation, or 12% less died (HR 0.58; 90%CI 0.33-1.00). Only the most recent and largest RCT by Salama et al [13] showed a higher survival rate in patients treated with TCZ. In this study, 377 subjects with COVID-19 pneumonia, 64% requiring low-flow and 26% on non-invasive and high flow oxygen supplementation, were randomized to TCZ (n=259, 8mg/kg, one or two doses) or placebo. By day 28 HR for mechanical ventilation or death was 0.56 (95%CI: 0.33-0.97, P=0.04), while still the number of patients who died by that day of any reason was 10.4% in TCZ-group vs 8.6% in the placebo group.

Summarizing, all aforementioned randomized clinical trials despite a rather homogenous population included, which is patients with COVID-19 pneumonia mainly requiring oxygen supplementation but not ventilated, it was not possible to visualize the obvious survival benefit of TCZ. Similarly in our study, the overall mortality was comparable between TCZ and non-TCZ patients (odds ratio, OR 1.05; 95%CI: 0.61-1.80). Furthermore, the group receiving TCZ showed even lesser odds of clinical improvement after 14 and 21 days of therapy (OR 0.61 and 0.64, respectively), while it was comparable to the standard of care therapy at the end of observation, ie. after 28 days (OR 0.87). On the other hand, only analyses in specific subgroups showed not only a higher survival rate but also more rapid clinical improvement in patients treated with TCZ. It was quite striking that in previous studies mortality HR had quite outsized confidence intervals, suggesting another factor playing the predictive role of TCZ efficacy. Surprisingly enough, cited above RCT did not evaluate baseline IL-6 serum concentration even since TCZ is aimed at blocking an IL-6 proinflammatory pathway. Indeed, we performed a detailed subgroup analysis aiming at the development of predictors of TCZ-response in COVID-19 subjects. Not unpredictably, the best response to TCZ concerning decreasing 28-day mortality (OR=0.27; 95%CI: 0.10-0.78, 11% vs 31%, P=0.02) was observed in subjects with baseline serum IL-6>100 pg/mL, while it was not observed in subject with baseline IL-6 50-100 pg/mL and below 50 pg/mL. Importantly, an IL-6 level of more than 100pg/mL was observed in appr. 18% of all studied patients, possibly explain why the difference was not noted in overall studied groups in the aforementioned RCT but also our study. This observation also underlines the pathogenetic complexity of cytokine imbalance during COVID-19. It is known that cytokine storm in

COVID-19 consists of various, not necessarily overlapping, soluble immune mediators (SIMs) including IL-1β, IL-6, IL-8, tumor necrosis factor-alpha (TNF-α) which could yield different predictive value [20]. Interestingly, Mathew et al [21] in their elegant study shown at least three different immunotypes of COVID-19, 1-3 depending on the cluster of differentiation (CD)4+ cell, CD8+ cell, B-cell, and plasmablasts activation/exhaustion, which was associated with different outcomes but also most likely with different levels of cytokines. Interestingly, despite baseline IL-6 in our study TCZ administration was not associated with baseline CRP, D-dimer, or lymphocyte concentrations, which are also regarded as factors associated with prognosis [22].

Another important finding of our study was that the highest reduction in mortality, the need for mechanical ventilation, and best clinical improvement at day 28 in patients receiving TCZ vs standard of care (SOC)-therapy was observed in patients with baseline IL-6>100ng/mL and Sp02<90% (11 vs 50%, 5 vs 29%, 75 vs 37% respectively), which was not the case in subjects with Sp02≥90%. This observation might further underline that in subjects with severe hypoxia further deregulation between IL-6 levels and other cytokines is present and possibly IL-6 activation is deeper and not counterbalanced by regulatory mechanism, which could explain why the effectiveness of TCZ is more significant. Also in our study correlation analyses showed the correlation pattern of IL-6 and some soluble immune mediators are different in patients with oxygen saturation lower and higher/equal to 90%.

The results of our study should be taken with some caution since its retrospective real-world evidence design and in some subgroup analyses, the number of participants is small. Moreover, some patients in both arms received additional medication, which could affect the outcome of the disease. On the other hand, patients receiving TCZ and non-TCZ SOC therapies seem to be well balanced with regard to comorbidities and co-medications for COVID-19, and undoubtedly its advantage is the assessment of baseline serum IL-6 concentration. Indeed, while only one another single-center study showed a beneficial effect of TCZ mainly in patients with higher IL-6 [23], our data in a real-world large dataset seems to guide the effective use of TCZ in COVID-19.

In conclusion, the possible benefit from the treatment of COVID-19 with tocilizumab can be achieved in selected subpopulations only. This regimen can be associated with reduced mortality and the need for mechanical ventilation in patients with a baseline concentration of interleukin 6 exceeding 100 pg/ml, particularly if they need oxygen supplementation due to oxygen saturation of ≤90%. Patients who worsened within the initial 7 days of hospitalization can also obtain some benefits from tocilizumab administration, but it should be clarified in further studies on a bigger number of patients.

## Data Availability

data will be available upon request from the first/corresponding author

## Authors contribution

All authors had full access to all the data in the study and take responsibility for its integrity and the accuracy of the analysis. RF was responsible for the study concept, design, acquisition of data, verification of the underlying data, and drafting the manuscript. JJ and JP were responsible for statistical analysis and interpretation of data and drafting some parts of the article. All other authors were responsible for the acquisition of data and revised the final content of the manuscript. All authors approved the final version of the article before submission.

## Acknowledgments

The study was supported by the Polish Association of Epidemiologists and Infectiologists and Medical Research Agency

